# Eccentric-focused rehabilitation promotes myelin plasticity in individuals with chronic, incomplete spinal cord injury

**DOI:** 10.1101/2020.04.27.20079970

**Authors:** Timothy D. Faw, Bimal Lakhani, Hanwen Liu, Huyen T. Nguyen, Petra Schmalbrock, Michael V. Knopp, Keith R. Lohse, John L.K. Kramer, Dana M. McTigue, Lara A. Boyd, D. Michele Basso

## Abstract

**Background:** Myelin plasticity has emerged as a novel mechanism by which the nervous system can change with experience, offering new potential for rehabilitation-induced recovery after neurotrauma. This first-in-human study investigated whether innovative, downhill locomotor rehabilitation promotes myelin plasticity in individuals with chronic, incomplete spinal cord injury (SCI).

**Methods:** Of 20 individuals with SCI that enrolled, 4 passed the imaging screen and had myelin water imaging (MWI) before and after a 12-week (3 times/week) downhill locomotor treadmill training program (SCI+DH). One individual was excluded for imaging artifacts. Uninjured control participants (n=7) had two MWI sessions within the same day. Changes in myelin water fraction (MWF), a histopathologically-validated myelin biomarker, were analyzed in *a priori* motor learning and non-motor learning brain regions and the cervical spinal cord using statistical approaches appropriate for small sample sizes.

**Results:** Within SCI+DH individuals, significantly more motor learning regions showed increased MWF than non-motor learning regions (p<.05). Compared to Control, MWF in the SCI+DH group increased in white matter underlying postcentral and precuneus cortices, combined motor learning brain regions, and ventral spinal cord (p<.05). To account for small sample size, an estimation-based approach showed the pattern of MWF increase was specific to training and region.

**Conclusion:** Downhill training increased MWF in brain regions specifically associated with motor learning and in the ventral spinal cord.

**Trial Registration:** ClincialTrials.gov (NCT02498548, NCT02821845)

**Funding:** National Institutes of Health [F31NS096921 (TDF), R21HD082808 (DMB)], Craig H. Neilsen Foundation [316282 (DMB)], Foundation for Physical Therapy Research [Promotion of Doctoral Studies Level II Scholarship (TDF)]

## Introduction

Experience-dependent plasticity has primarily focused on neuronal and gray matter changes (1-3). More recently, white matter plasticity emerged as a novel central nervous system (CNS) component of motor learning (4-10). Repetitive activation of neural pathways during motor skill training appears to trigger increased myelin (11). Specifically, activated neurons release neurotransmitters not only at the synaptic cleft, but also along the length of the axon (12, 13). These neurotransmitters then bind to receptors on oligodendrocyte progenitor cells causing their proliferation and differentiation into mature, myelinating oligodendrocytes (12-26). In fact, the molecular programs that lead to new oligodendrocytes and myelin formation are activated within hours of motor learning (9) and are ultimately required for motor learning to occur (8). Even small changes in myelination can greatly impact neural network function (27, 28), making it especially important in spinal cord injury (SCI) where myelin damage and sensorimotor dysfunction are hallmark features.

To date, most *in vivo* studies have used diffusion weighted imaging (DWI) to quantify training-induced myelin plasticity, but advances in imaging provides a neurobiologically-specific biomarker of myelin (4-7, 29-31). While DWI detects changes in the microstructural components of white matter (32), the metrics also reflect numerous factors not necessarily related to myelin (33). Alternatively, myelin water imaging (MWI) uses multicomponent T2 relaxation to generate a distribution of T2 proton relaxation times, which can be separated into three distinct components: 1) a short component (<40ms) that corresponds to water molecules between the myelin bilayers, 2) an intermediate component (∼80ms) attributed to intra/extracellular water, and 3) a long component (∼2s) corresponding to cerebral spinal fluid (29, 34-37). The *in vivo* measure of myelin, or myelin water fraction (MWF), is derived from the ratio of the short component to the total T2 distribution and demonstrates strong histopathological validity for multiple CNS regions, species, and myelin-related diseases (30, 31, 38-45). Despite its utility, only one study to date has utilized MWI to quantify motor learning-induced myelin plasticity in healthy individuals (10). No studies have used MWF as a biomarker of myelin plasticity in individuals with SCI or in the context of locomotor learning.

Intensive locomotor rehabilitation has shown the most promise for improving functional recovery after SCI, even in the chronic stage (46-53). Yet even after intensive rehabilitation, recovery is often incomplete and significant sensory and motor dysfunction persist (51). Thus, there is an acute need for novel approaches to facilitate motor learning and plasticity. Increasing task complexity produces greater motor learning-related change and relies on both neural and myelin plasticity (8, 54, 55). In the current study, we increased locomotor complexity by performing treadmill training on a downslope. Downhill walking relies almost entirely on eccentric, or lengthening, muscle contractions (56-58), and produces greater challenge to joint kinetics (58-60), balance and stability (61), and muscle activation (62, 63) as compared to walking on level surfaces. Eccentric muscle actions require greater neural control than concentric contractions, because precise integration between descending supraspinal motor signals and afferent information from the periphery must occur to achieve efficient motor performance (64). Eccentric training is especially compelling in the context of SCI given that it uniquely increases brain activation in order to inhibit the muscle spindle afferent drive from the stretched muscle to the motor neuron to produce controlled muscle lengthening (64-72). Indeed, downhill walking decreases spinal reflex excitability in healthy individuals (73) and those with multiple sclerosis (74, 75). Depressed spinal reflexes have in turn been linked to improved walking function after SCI (76, 77). As such, this downhill approach may be particularly suited to better and uniquely engage the CNS and may promote myelin plasticity after SCI. Interestingly, we showed that hindlimb electromyography is normalized during downhill walking in rats with SCI (63). It is unknown whether experience-dependent myelin plasticity occurs after SCI or whether rehabilitation paradigms can promote myelin plasticity in the chronic phase of injury. Therefore, the purpose of this first-in-human study was to investigate the longitudinal changes in brain and spinal cord MWF after a 12-week eccentric-focused locomotor training program in individuals with chronic, incomplete SCI.

## Methods

### Study Approval

This myelin imaging study recruited participants from two ongoing clinical trials designed to investigate the effect of eccentric-focused, downhill training on motor control after SCI (NCT02498548, NCT02821845). All individuals provided informed consent in accordance with the Declaration of Helsinki and all aspects of the study were approved by The Ohio State University’s (OSU) Institutional Review Board.

### Participants

Participants with SCI (n=20) were recruited from the primary study and met the following inclusion criteria: age 18 – 85 years old, motor incomplete SCI within C1 – T10, some capacity to step over ground and on a treadmill, had completed an intensive treadmill-based locomotor training program, and were at least 6 months since discharge from outpatient rehabilitation. Exclusion criteria were: lower motor neuron injury innervating the legs, co-existing neurological conditions (i.e. brain injury), cognitive conditions that prevented informed consent, pregnancy, active deep vein thrombosis, skin wounds in regions where harness or hands provide support, or a history of Botox use for spasticity management within the 3 months prior to enrollment. Uninjured control participants (n=7) were recruited from the primary study and the community. All participants completed MRI safety screening. The disposition of participants and reasons for exclusion is shown in Figure 1. See Table 1 for demographics of participants who completed the study and all data processing.

**Table 1.** Demographics and clinical characteristics of participants who completed data processing.

| ID | Age,<br>years | Sex<sup>a</sup> | BMI | Race | Ethnicity<br>(Hispanic/Latino) | Cause of<br>Injury | Level<br>of<br>SCI | Time<br>Since<br>Injury,<br>years | AIS<br>Classification | Assistive<br>Device |
| --- | --- | --- | --- | --- | --- | --- | --- | --- | --- | --- |
| SCI-01 | 35 | F | 24.2 | White | No | Epidural<br>Abscess | T6 | 6.25 | D | Rollator |
| SCI-02 | 64 | M | 27.9 | Black or<br>African American | Yes | Traumatic | C1 | 48 | D | Wheelchair |
| SCI-03 | 63 | F | 23.3 | White | No | Infection | T9 | 42 | D | Wheelchair |
| **Average** | **54**<br>**(16.5)** | - | **25.1**<br>**(2.4)** | - | - | - | - | - | - | - |
| Control-01 | 35 | M | 25.3 | White | No | - | - | - | - | - |
| Control-02 | 66 | M | 24.7 | White | No | - | - | - | - | - |
| Control-03 | 64 | F | 25.8 | White | No | - | - | - | - | - |
| Control-04 | 28 | F | 27.1 | White | Yes | - | - | - | - | - |
| Control-05 | 59 | M | 25.8 | White | No | - | - | - | - | - |
| Control-06 | 34 | F | 22.0 | White | Yes | - | - | - | - | - |
| Control-07 | 36 | F | 23.0 | White | No | - | - | - | - | - |
| **Average** | **46**<br>**(16.2)** | - | **24.8**<br>**(1.8)** | - | - | - | - | - | - | - |
| **p-Value** | .5583 | 1.0 | .9667 | .3000 | 1.0 | - | - | - | - | - |

<sup>a</sup> Ascertained by self-report. Average values are expressed as mean (SD). Continuous variables are compared using the Mann-Whitney U nonparametric test. Categorical values are compared using Fisher's Exact Test. Abbreviations: AIS, ASIA Impairment Scale; BMI, Body Mass Index; C, Cervical; F, Female; ID, Participant Identifier; M, Male; SCI, Spinal Cord Injury; T, Thoracic

**Figure 1.**
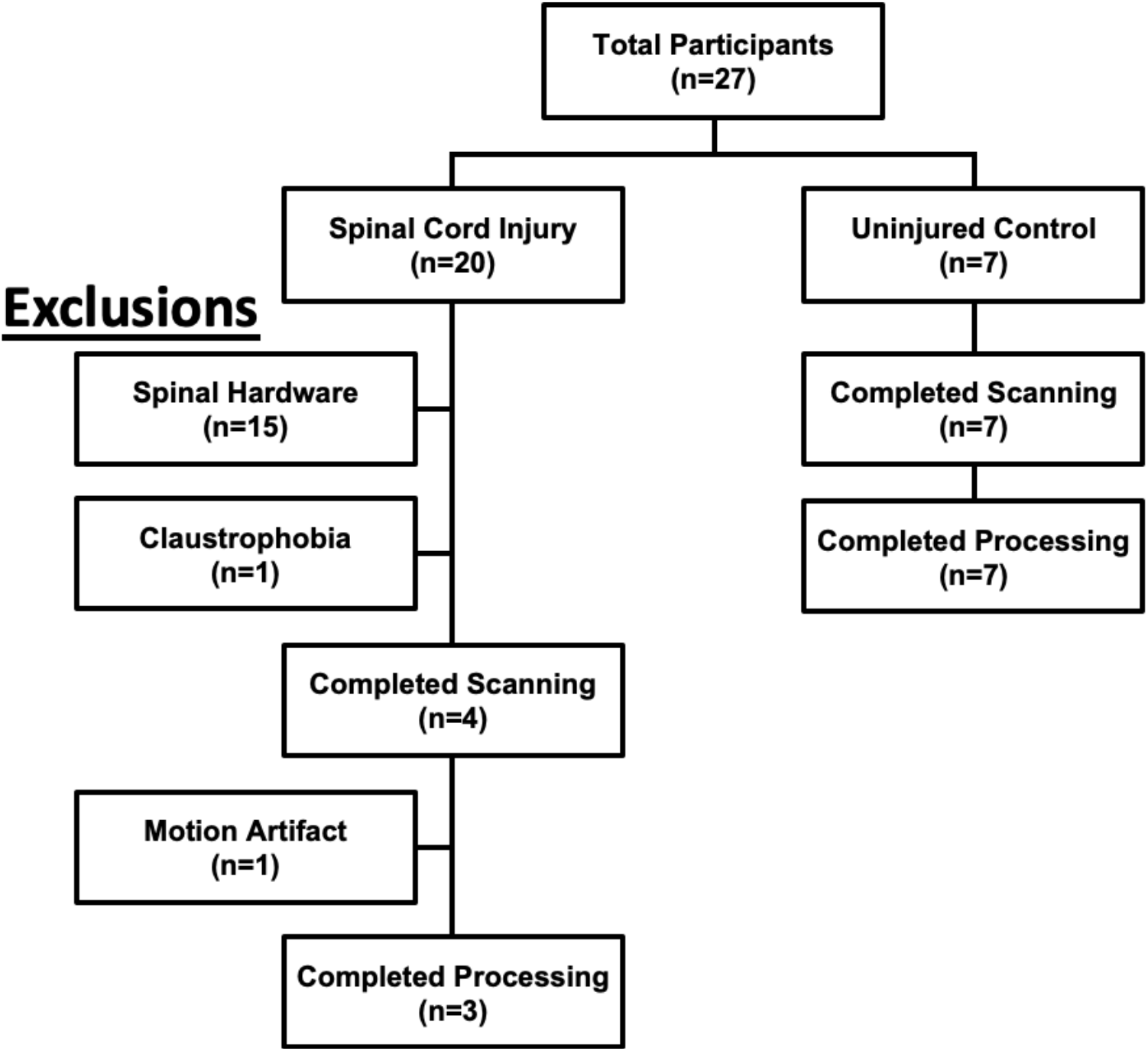
*Disposition of participants*. The timing and reason for exclusions from the MRI study are noted on the left.

### Eccentric-focused Downhill Training Intervention

The downhill training intervention for participants with SCI began at least 6 months after neurorehabilitation ended (Figure 2). Trainer-assisted downhill stepping occurred 3 times a week for 12 weeks on a 10% grade, using an instrumented, split-belt treadmill (Bertec; Columbus, OH) with custom body-weight support system. Training duration totaled 20 minutes per session, performed in up to 5-minute stepping bouts separated by at least three, 5-minute seated rests. This study design was used as equivalent rest periods can reduce or eliminate delayed onset muscle soreness from downhill eccentric exercise (78, 79). Individualized eccentric training complexity occurred by modulating BWS and treadmill walking speed to target frontal plane hip motion or sagittal plane knee motion (80). A team led by a licensed physical therapist in collaboration with up to three clinical rehabilitation assistants provided intermittent manual assistance to allow error-based motor learning. The objective was self-correction by participants with SCI to attain normal kinematic alignment of the head, trunk, pelvis and legs. Trainers intervened briefly when biomechanical stepping errors predominated. Verbal cues during stepping bouts promoted knowledge of performance, and verbal feedback during rest breaks provided knowledge of results. Verbal cues were structured to facilitate external focus of attention when possible.

**Figure 2.**
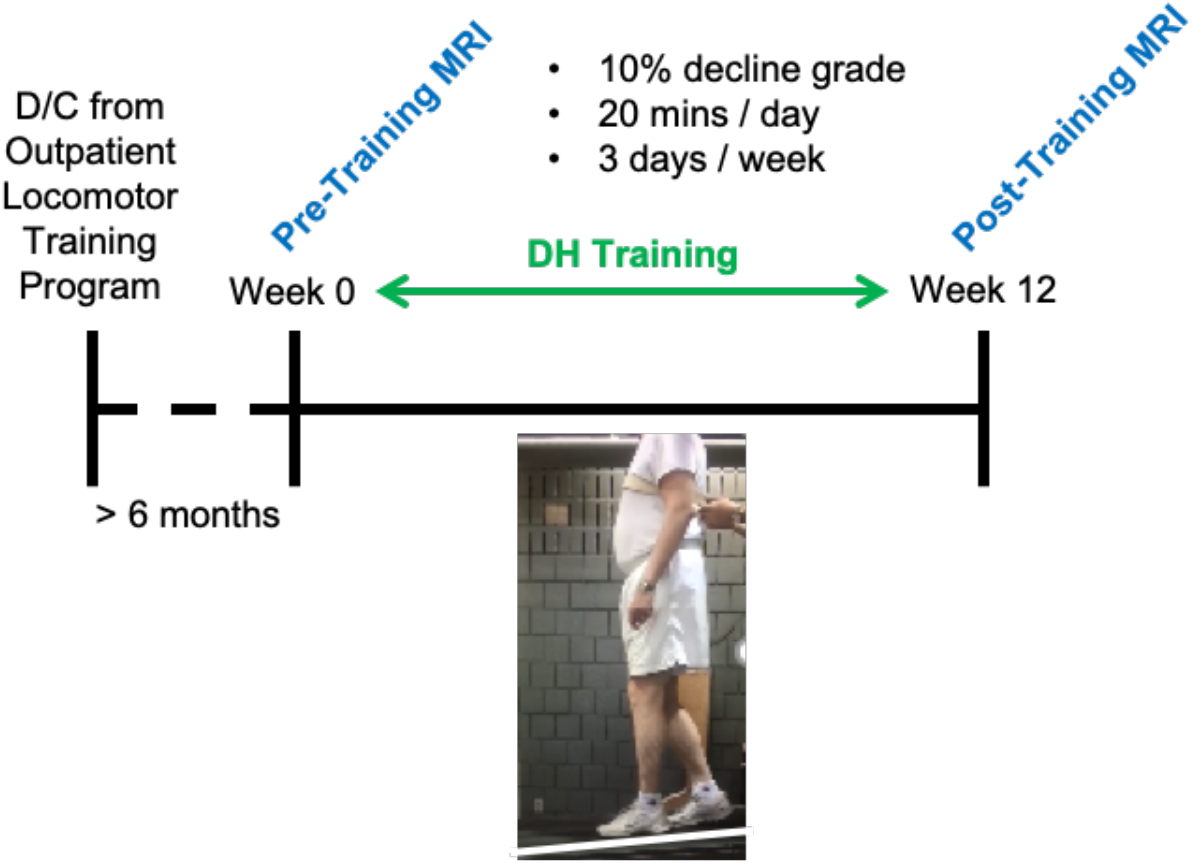
*Experimental timeline*. Individuals with incomplete SCI completed the experimental timeline described above.

### MRI Acquisition

Magnetic resonance (MR) acquisition of the whole brain was performed at the OSU Wright Center for Innovation using a 3-tesla (T) MRI system (Ingenia CX, Philips Healthcare). To acquire images, a dStream head/spine radiofrequency (RF) coil was used. The following scans were collected of the brain: 1) High-resolution 3D fast field echo (FFE) T1-weighted sequence (repetition time (TR) = 9.9ms, echo time (TE) = 4.6ms, matrix = 232 × 192, in-plane field of view (FOV) = 232 × 232mm, slice thickness = 1mm, number of slices = 150, number of signal averages = 1, flip angle = 8°, turbo field echo (TFE) factor = 192, scan time = 5min 20sec) and 2) 32-echo gradient and spin echo (GRASE) T2-weighted sequence (TR = 1200ms, TE = 10,20,30…,320ms, flip angle = 90°, refocusing flip angle = 180°, matrix = 224 × 90, in-plane FOV = 224 × 180mm, slice thickness = 1.5mm, number of slices = 80, number of signal averages = 1, turbo spin echo factor = 32, scan time = 9min 30sec).

To acquire images of the cervical spinal cord (C2-C4), the following scans were collected: 1) merged fast-field echo (M-FFE) T1-weighted sequence (TR = 650ms, first TE = 4.1ms, echo spacing = 7ms, matrix = 188 × 188, in-plane FOV = 150 × 150mm, slice thickness = 2.5mm, number of slices = 16, number of signal averages = 2, flip angle = 28°, scan time = 4min 10sec) and 2) 32-echo GRASE T2-weighted sequence (TR = 1500ms, TE = n*10ms, flip angle = 90°, refocusing flip angle = 180°, matrix = 240 × 196, in-plane FOV = 180 × 147mm, slice thickness = 2.5mm, number of slices = 16, number of signal averages = 1, TSE factor = 32, scan time = 9min 10sec).

For individuals with SCI, MRI-acquisition occurred within 6 weeks prior to downhill training (range = 3d to 39d) and within 24 hours of the final training session. For test-retest reliability in uninjured control participants, scans of the brain and spinal cord were performed back-to-back, separated by changing of RF coils. The participants were then allowed to adjust positioning before the retest condition.

### Generation of MWF Maps

Generation of brain and spinal cord MWF maps from T2 relaxation imaging used the latest non-negative least squares method (35), implemented with an extended phase graph algorithm to correct for the stimulated echoes (29). The T2 distribution of each imaging voxel was partitioned into short (<40ms), intermediate (40-200ms), and long (>1500ms) components using an in-house MATLAB code (MATLAB R2013b, MathWorks Inc) developed by collaborators at the University of British Columbia (code can be requested from https://mriresearch.med.ubc.ca/news-projects/myelin-water-fraction/). In this method, the short component corresponds to water molecules between myelin bilayers (35, 81). The MWF was defined as the sum of the amplitudes within the short T2 component divided by the amplitude sum of the total T2 distribution. Voxel-based MWF maps were generated for each participant.

### Processing of Brain Imaging

Further processing of brain MR data was performed as previously described by Lakhani et al (82). Here, we utilized the freely available FreeSurfer Software Suite v5.3 (https://surfer.nmr.mgh.harvard.edu/) to perform all morphometric procedures, including automated cortical reconstruction and volumetric segmentation. The technical details of these procedures have been described previously (82-92). Briefly, this processing included motion correction and averaging (90) of multiple volumetric T1 weighted images, removal of non-brain tissue using a hybrid watershed/surface deformation procedure (88), automated Talairach transformation, segmentation of the subcortical white matter and deep gray matter volumetric structures (including hippocampus, amygdala, caudate, putamen, ventricles) (86, 87), intensity normalization (93), tessellation of the gray matter white matter boundary, automated topology correction (85, 94), and surface deformation following intensity gradients to optimally place the gray/white and gray/cerebrospinal fluid borders at the location where the greatest shift in intensity defines the transition to the other tissue class (83, 84, 95). Images were automatically processed with the longitudinal stream (91). Specifically an unbiased within-subject template space and image was created using robust, inverse consistent registration (90). Several processing steps, such as skull stripping, Talairach transforms, atlas registration as well as spherical surface maps and parcellations were then initialized with common information from the within-subject template, significantly increasing reliability and statistical power (91). Visual inspection of FreeSurfer surface reconstructions occurred at each stage of the longitudinal processing pipeline. Manual correction of errors included removing dura misidentified as cortex and re-processing the current and subsequent steps.

Linear co-registration occurred in three steps, using established methods by Lakhani et al (82) and Deoni et al (92): 1) registration of the first echo from the T2 scan to the MWF map, 2) registration of the first echo of the T2 scan to the parcellated T1 anatomy, and 3) registration of the MWF map to the parcellated anatomy. Importantly, all processing occurred on the same workstation, using the same software version (96) and individualized intensity normalization was used to increase registration accuracy (93). The FreeSurfer-generated white matter parcellation map (89) was used to extract mean MWF from a priori identified regions of interest (ROI).

### Processing of Spinal Cord Imaging

Post-processing of spinal cord MR data was performed as described by Liu et al (97). Spinal cord segmentation of the GRASE image occurred using the Spinal Cord Toolbox PropSeg tool (98, 99), which generated a mask of the whole cord. For segmentation, we used the 16^th^ echo of the GRASE image because it has the greatest contrast between the spinal cord and cerebrospinal fluid. Visual inspections occurred and manual corrections to the mask were made when needed. The whole cord mask was then registered to the PAM50 template (100) using a nonlinear transformation that involved a combination of center of mass rotation, affine, symmetric normalization, and bsplinesyn algorithms. This process obtained the warping functions used to transform the GRASE images and subsequently MWF maps into the template space. Visual inspection of all registered images occurred. MWF values were extracted from the PAM50 atlas labels.

### Analysis and Statistics

For analysis of brain MWF data, subcortical white matter regions corresponding to areas known to be responsive to motor learning were identified *a priori*. Motor learning areas included white matter underlying the precentral, postcentral, paracentral, caudal middle frontal, superior frontal, inferior parietal, and precuneus cortices, and the anterior and posterior regions of the corpus callosum (CC) (101). Regions not expected to change following an eccentric-focused locomotor task were also identified *a priori*. Non-motor learning brain regions included white matter underlying the fusiform, inferior temporal, lateral occipital, middle temporal, superior temporal, lateral orbitofrontal, lateral occipital, and lingual cortices (101). Due to the bilateral nature of the locomotor task, MWF data from right and left ROI’s were combined and averaged to calculate on overall MWF value for each region. Within-session test-retest reliability was calculated from all uninjured control participants for all motor learning regions and three out of the seven non-motor learning regions (lateral occipital, lateral orbitofrontal, and lingual). Missing temporal lobe imaging data was identified on the GRASE scan of three healthy control individuals that affected the fusiform, inferior temporal, middle temporal, and superior temporal regions. Thus, test-retest reliability was determined in these regions using data from n=4 uninjured control participants without data loss. The intraclass correlation coefficient (ICC) was determined in all ROI’s. Regions with poor reliability, based on ICC values <0.5, were excluded from subsequent analyses (102).

To determine whether individual MWF changes could be interpreted as exceeding measurement error (103), we calculated the minimal detectable change (MDC) for each ROI using the formula MDC = SD * √ (1 – r) * 1.96 as described by Forrest et al (104). Here, SD represents the standard deviation of the difference between pre- and post-intervention measurements, r represents the ICC, and 1.96 is the 97.5% quantile of the standard normal distribution of a two-tailed test with the significance set at .05. Fisher’s exact test was used to determine whether motor learning regions were more or less likely to demonstrate increases exceeding the MDC.

For cohort analyses, we compared the mean percent change of the downhill trained SCI participants (SCI+DH) with the mean percent change of the test-retest data in the uninjured control group (Control) using a Mann Whitney U, nonparametric test to determine whether the mean percent MWF change within each ROI for the SCI+DH group exceeded the variability of the measure established by the Control group. Further, to account for the small number of subjects with SCI, we employed an estimation-based method of analysis that focuses on effect size. Using the 95% confidence intervals (CI), the mean percent change within *a priori* defined motor learning regions were contrasted against two sets of control regions: non-motor regions within the same SCI participant (within group) and homologous motor regions in uninjured control participants (between group). Change data from each ROI were pooled to determine an overall percent change for motor learning and non-motor learning regions. Here, we calculated a weighted mean and 95% CI based on the number of data points each individual contributed to the average to account for missing data from three uninjured control participants.

## Results

### Reliability of MWF Measures

In past research, myelin water imaging showed significant variation in MWF values across control white matter regions (105), but had good reliability for test-retest (10, 106-109), between vendors (110) and facilities (111), and across field strengths (112). To determine the stability of MWF values at our OSU facility, we measured the within-session test-retest reliability of the MWF signal in uninjured control participants. Two motor learning regions (paracentral and superior frontal) had poor single-session test-retest reliability and were excluded from further analyses. Moderate to excellent reliability of MWF occurred for remaining ROIs (ICC range: 0.503-0.907; Table 2). Spinal cord ROIs had good to excellent test-retest reliability of MWF values (ICC range: 0.814-0.917; Table 2).

**Table 2.** Within-session test-retest MWF reliability.

|  | **First MWF** | **Second MWF** | **ICC** |
| --- | --- | --- | --- |
| **<u>Brain Regions (Motor Learning)</u>** |  |  |  |
| Anterior Corpus Callosum | 0.095 | 0.095 | 0.907 |
| Posterior Corpus Callosum | 0.127 | 0.121 | 0.738 |
| Paracentral | 0.105 | 0.092 | 0.251 |
| Precentral | 0.092 | 0.092 | 0.577 |
| Postcentral | 0.076 | 0.075 | 0.801 |
| Caudal Middle Frontal | 0.056 | 0.055 | 0.807 |
| Superior Frontal | 0.039 | 0.035 | 0.342 |
| Inferior Parietal | 0.058 | 0.060 | 0.742 |
| Precuneus | 0.075 | 0.071 | 0.834 |
| **<u>Brain Regions (Non-Motor Learning)</u>** |  |  |  |
| Lingual | 0.066 | 0.069 | 0.709 |
| Lateral Orbitofrontal | 0.039 | 0.040 | 0.880 |
| Lateral Occipital | 0.074 | 0.075 | 0.795 |
| Fusiform<sup>a</sup> | 0.062 | 0.060 | 0.857 |
| Inferior Temporal<sup>a</sup> | 0.058 | 0.057 | 0.658 |
| Middle Temporal<sup>a</sup> | 0.060 | 0.058 | 0.503 |
| Superior Temporal<sup>a</sup> | 0.066 | 0.062 | 0.551 |
| **<u>Spinal Cord Regions</u>** |  |  |  |
| Whole Cord | 0.219 | 0.220 | 0.898 |
| Dorsal Columns | 0.259 | 0.261 | 0.917 |
| Lateral Funiculi | 0.226 | 0.229 | 0.814 |
| Ventral Funiculi | 0.218 | 0.212 | 0.863 |

<sup>a</sup> n=4 participants. Abbreviations: ICC, Intraclass Correlation Coefficient; MWF, Myelin Water Fraction

### Minimal Detectable Change between MWF Measures

To determine whether individual, pre-post changes in MWF were likely to exceed measurement error (104), we calculated the MDC. In brain regions, MDCs ranged from 0.0032 in the middle temporal region to 0.0122 in the posterior corpus callosum (Table 3). Greater MDC variability occurred in spinal cord regions, ranging from 0.0028 in the whole cord to 0.0389 in the lateral funiculi (Table 3).

**Table 3.** Minimal detectable change of MWF values.

|  | **<u>MDC</u>** |
| --- | --- |
| **<u>Brain Regions (Motor Learning)</u>** |  |
| Anterior Corpus Callosum | 0.0029 |
| Posterior Corpus Callosum | 0.0122 |
| Precentral | 0.0117 |
| Postcentral | 0.0064 |
| Caudal Middle Frontal | 0.0062 |
| Inferior Parietal | 0.0041 |
| Precuneus | 0.0082 |
| **<u>Brain Regions (Non-Motor Learning)</u>** |  |
| Lingual | 0.0149 |
| Lateral Orbitofrontal | 0.0045 |
| Lateral Occipital | 0.0068 |
| Fusiform | 0.0109 |
| Inferior Temporal | 0.0028 |
| Middle Temporal | 0.0032 |
| Superior Temporal | 0.0040 |
| **<u>Spinal Cord Regions</u>** |  |
| Whole Cord | 0.0028 |
| Dorsal Columns | 0.0313 |
| Lateral Funiculi | 0.0389 |
| Ventral Funiculi | 0.0114 |

Abbreviations: MWF, Myelin Water Fraction; MDC, Minimal Detectable Change

### Downhill Training Increases Brain MWF in Individuals with Chronic SCI

Individual responses to downhill training were determined by comparing pre-training MWF values in motor learning and non-motor brain regions with those post-training. Participant SCI-01 had greater MWF after downhill training that exceeded the MDC in 6 out of 7 motor learning regions, with several regional increases being substantial and the smallest remaining above 10% (Fig. 3). Substantial increases of 26% for sensory, 33% for motor and 40% in caudal middle frontal regions were observed. Conversely, 1 non-motor learning region showed an increase and 1 showed a decrease in MWF that exceeded the MDC after training.

**Figure 3.**
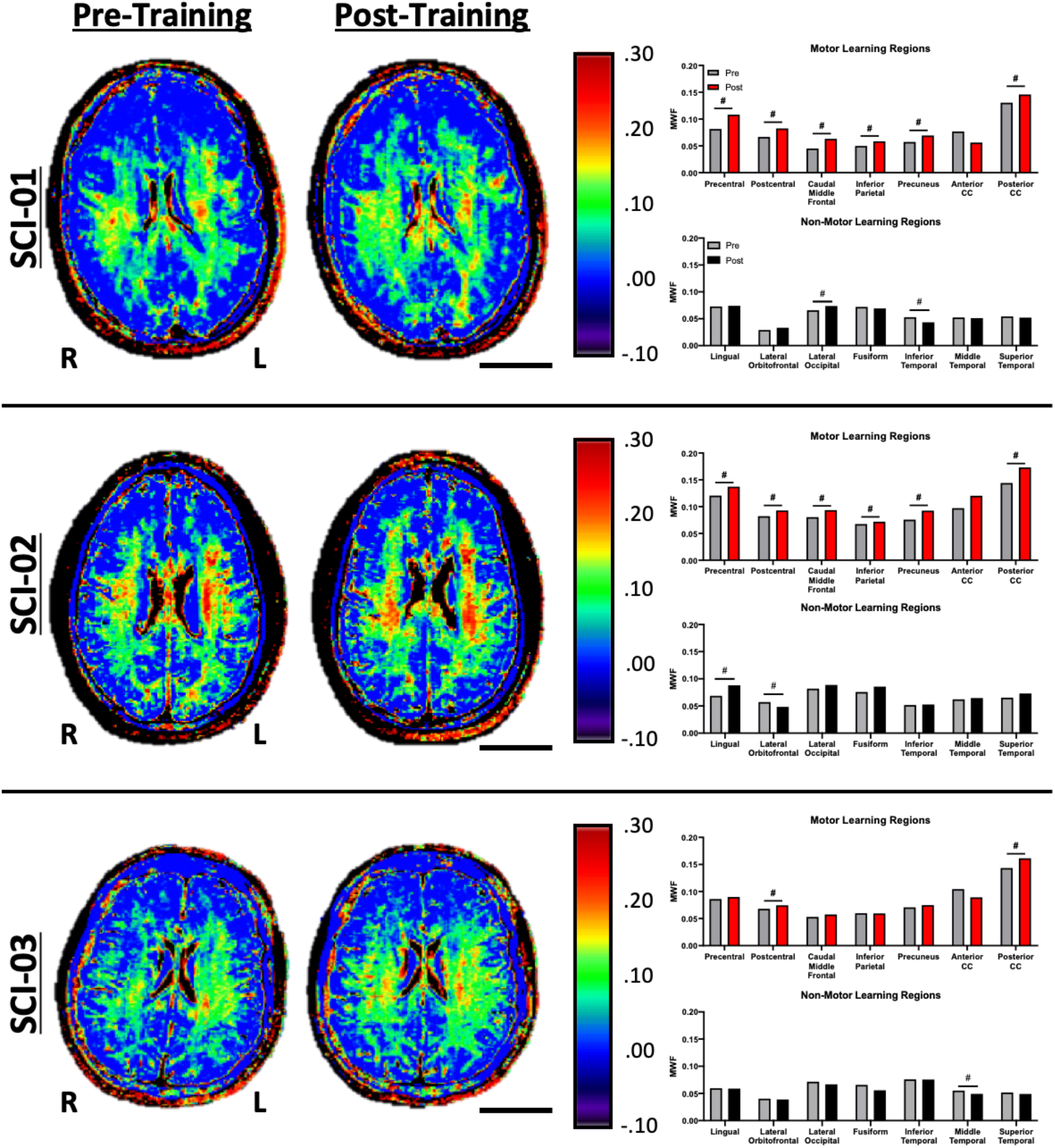
*Downhill training increases brain MWF in individuals with chronic SCI*. Pre- and post-downhill training MWF maps are shown from three individuals with SCI. Scale bar = 5cm. Quantifications of MWF within region of interest are shown. # = individual change exceeded the minimal detectable change (MDC) for that region.

Participant SCI-02 had MWF increases exceeding the MDC across 6 out of 7 brain motor learning regions after downhill training (Fig. 3). The largest increase occurred in the anterior corpus callosum (24%), followed by precuneus (22%) and posterior corpus callosum (20%). In non-motor learning regions for SCI-02, MWF increased (lingual region) and decreased (lateral orbitofrontal region) beyond MDC.

Participant SCI-03 showed the fewest brain changes in response to downhill training (Fig. 3). For this participant, MWF increased in 2 of 7 motor learning regions and decreased in 1 of 7 non-motor learning regions relative to MDC after training. The greatest increase in MWF occurred in the posterior corpus callosum (12%), followed by postcentral (10%).

A Fisher’s exact test demonstrated that significantly more brain regions associated with motor learning showed increases in MWF in our group of SCI participants as compared to non-motor learning areas. (Fig. 4; p=.0003).

**Figure 4.**
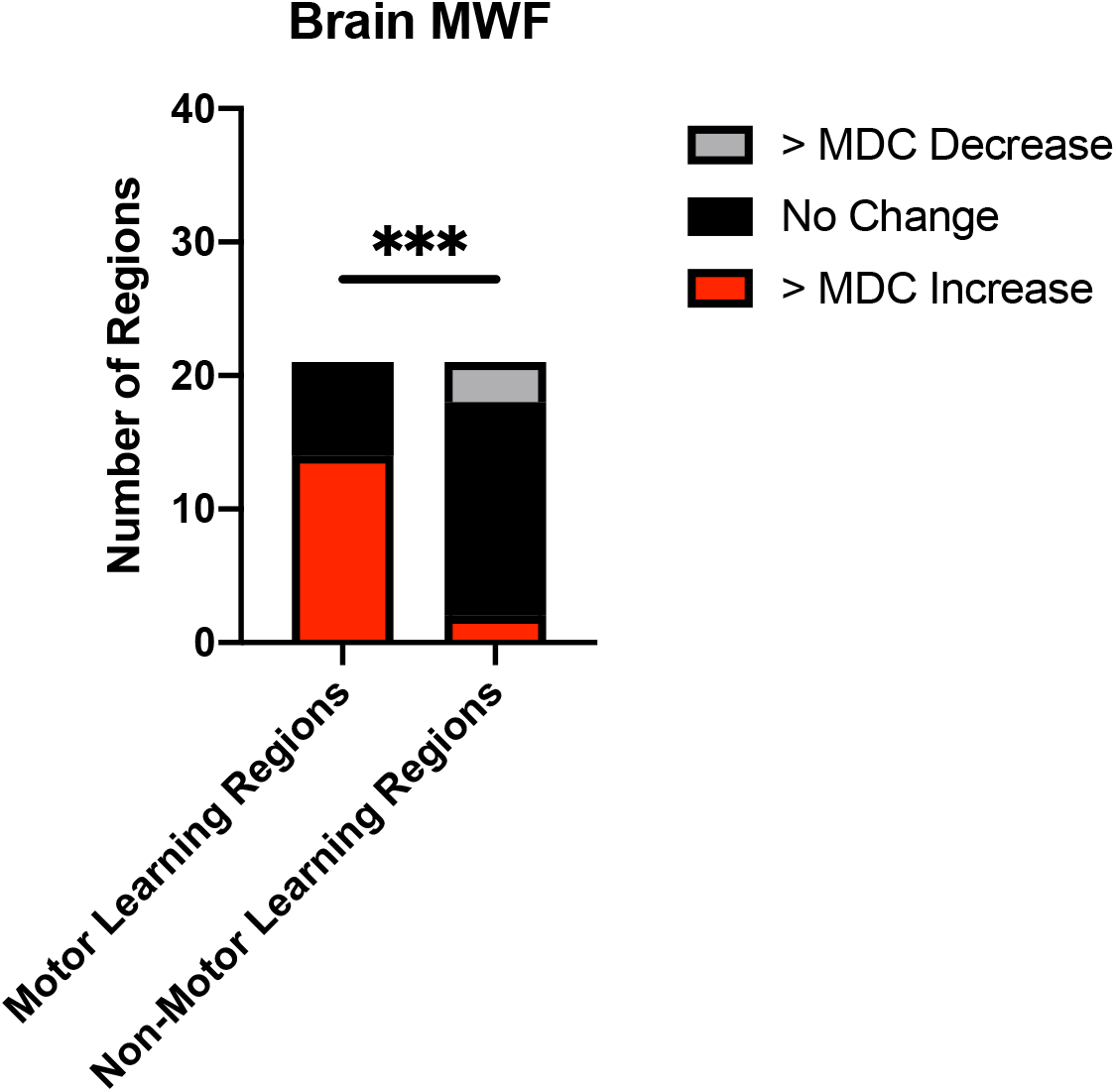
*MWF is more likely to increase in brain motor learning regions after downhill training*. Quantification of individual MWF changes between motor learning and non-motor learning regions of individuals with SCI. Comparison between the number of regions demonstrating MWF increase above MDC (> MDC increase, red) and those with either no change (black) or decreased MWF (< MDC decrease, gray). Significantly more motor learning regions had MWF increase beyond MDC than non-motor learning regions (***p<.001; Fisher’s Exact Test).

### Downhill Training Promotes Myelin Plasticity in Brain Motor Learning Regions

To determine whether group MWF changes exceeded measurement variability, we compared the average percent change between pre- and post-training values in the SCI+DH group with the test-retest percent change values in uninjured control participants (Fig. 5). The SCI+DH group had significantly greater MWF than Control in 2 of 7 motor learning regions (postcentral, p=.0167; and precuneus, p=.0167). Similarly, the SCI+DH group showed greater combined percent change across all motor learning regions as compared to non-motor learning areas (p=.033). No between-group differences in MWF occurred in any of the non-motor learning regions (p>.05).

**Figure 5.**
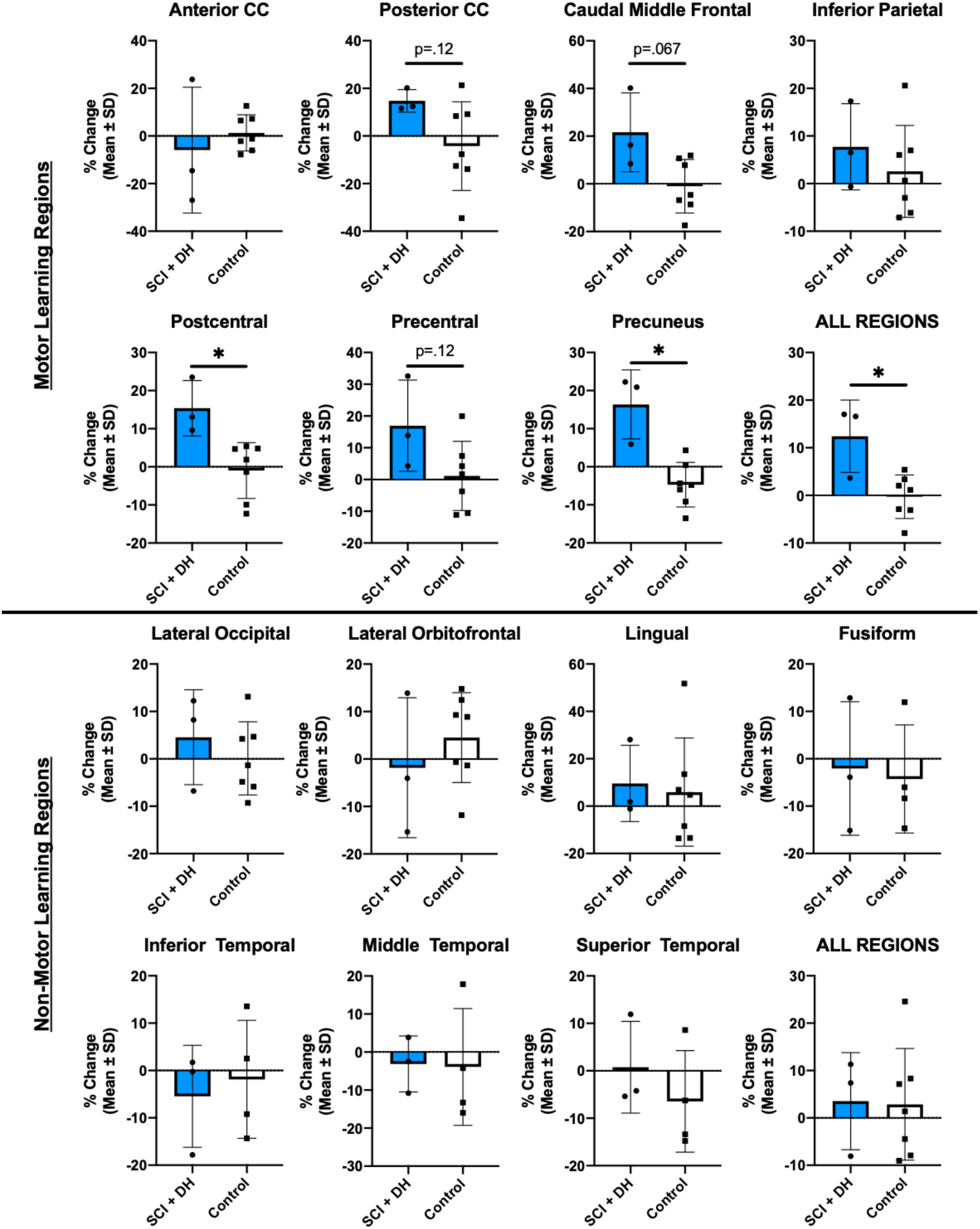
*Changes in Brain MWF by Region of Interest*. Between-group comparison of percent MWF change by region of interest associated with motor learning and non-motor learning areas. Graphs with * indicate p<.05 between SCI+DH and Control groups (Mann-Whitney U Test).

### Downhill Training Increases Ventral Funiculus MWF in Individuals with SCI

Individual white matter plasticity responses in the cervical spinal cord were also identified by comparing pre- and post-training MWF values in major white matter funiculi (Fig. 6). The MWF tended to decrease in the dorsal columns of all three participants and exceeded the MDC for SCI-01. Conversely, MWF increased in the ventral funiculi of all three participants and in 2 of 3 participants exceeded the MDC. Individual MWF changes in the lateral funiculi and the whole spinal cord failed to surpass the MDC.

**Figure 6.**
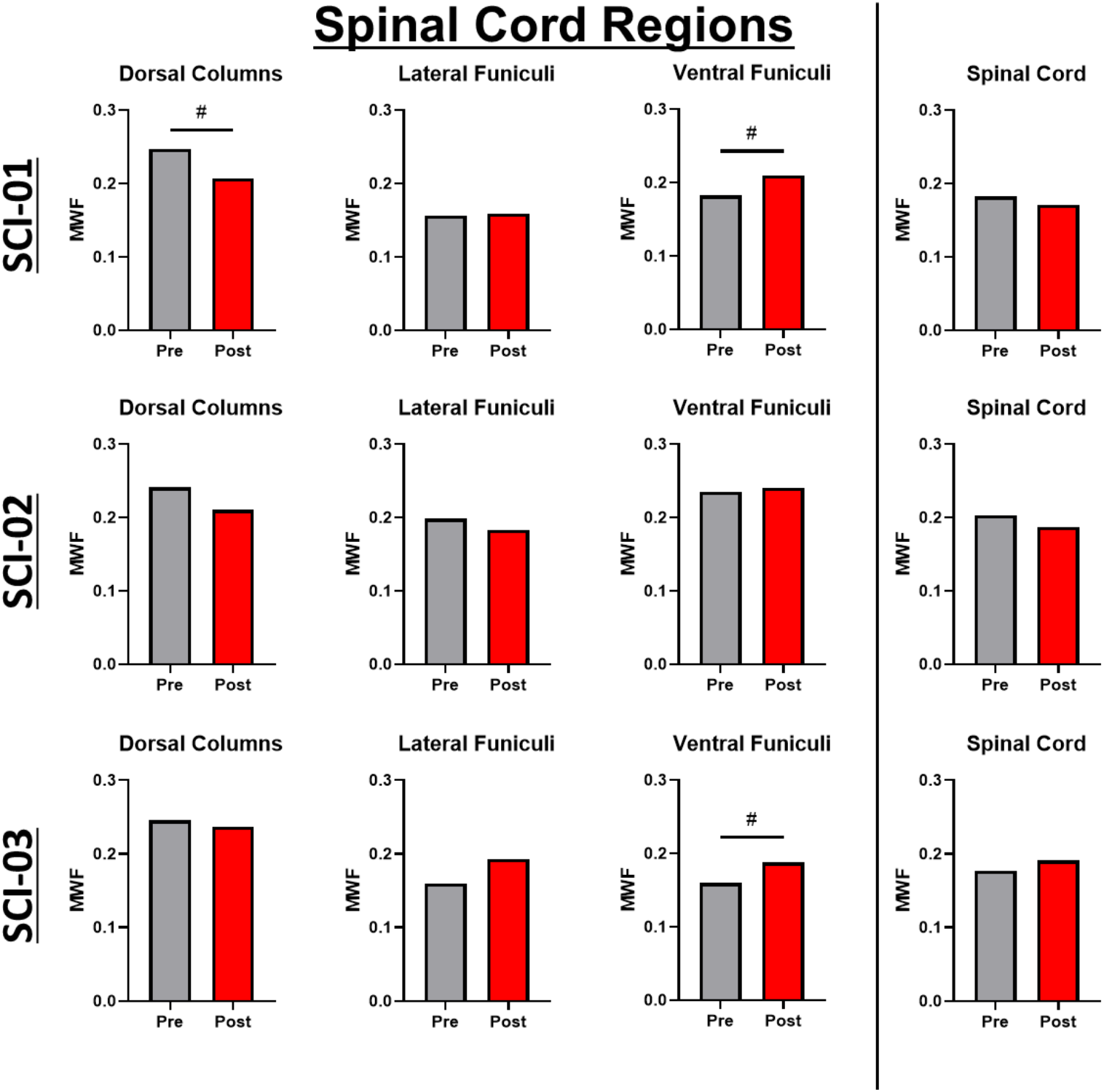
*Individual spinal cord MWF*. Quantifications of pre- and post-training MWF by white matter region and whole spinal cord at C2 are shown for each individual. Regions with # indicate that the MWF change exceeded the MDC for that region.

We compared the mean percent change in values between the SCI+DH cohort and test-retest percent change scores for the Control group for all spinal cord ROI’s (Fig. 7). The MWF increase in the ventral funiculi of SCI individuals post downhill was significantly greater than measurement variability of the control group (p=.033) but the decrease of MWF in the dorsal columns was not different between groups.

**Figure 7.**
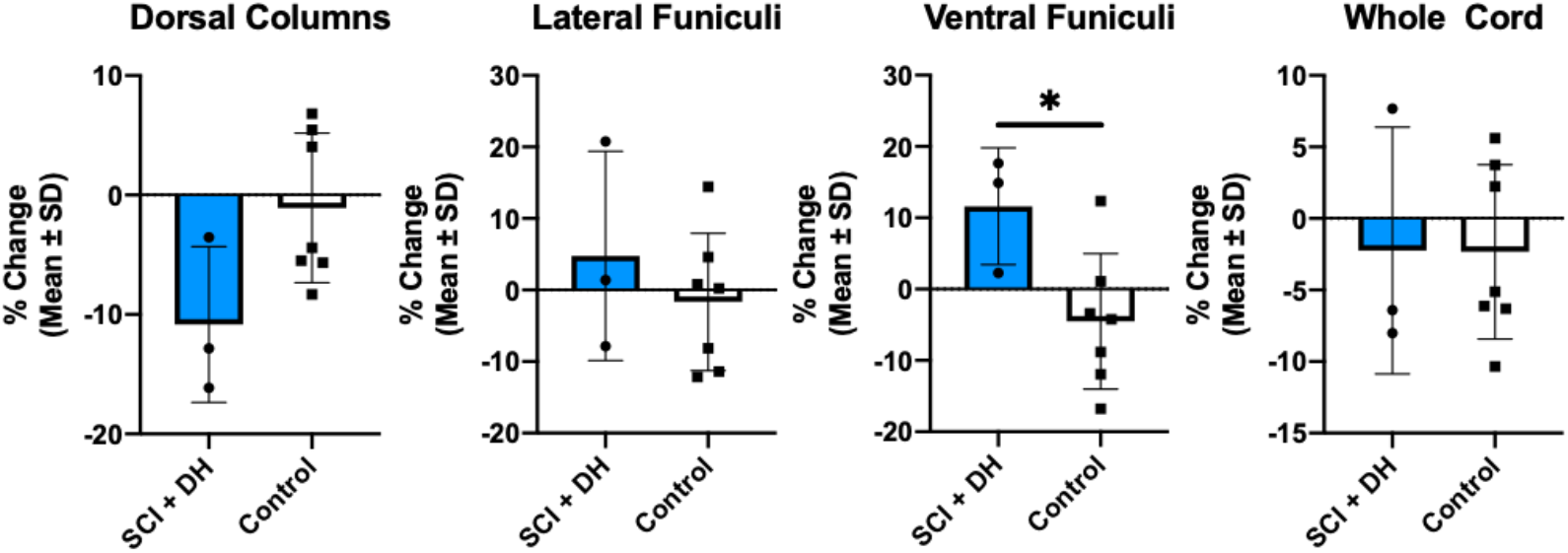
*Changes in Spinal Cord MWF by Region*. Between-group comparison of percent MWF change by white matter region and for the whole cervical spinal cord at C2. Graphs with * indicate p<.05 between SCI+DH and Control groups (Mann-Whitney U Test).

### Myelin Plasticity is Specific to Training and Region

To account for the small number of subjects with SCI, we used an estimation-based approach to determine effects of downhill training on myelin plasticity in the brain and spinal cord. In the brain, there was a positive, rightward shift in MWF CI for 6 of 7 motor learning regions following downhill training. Clustering at the zero line for MWF values in non-motor regions within individuals with SCI and in motor learning and non-motor learning regions of uninjured control participants (Fig. 8) indicate that downhill training did not induce generalized brain myelin responses.

**Figure 8.**
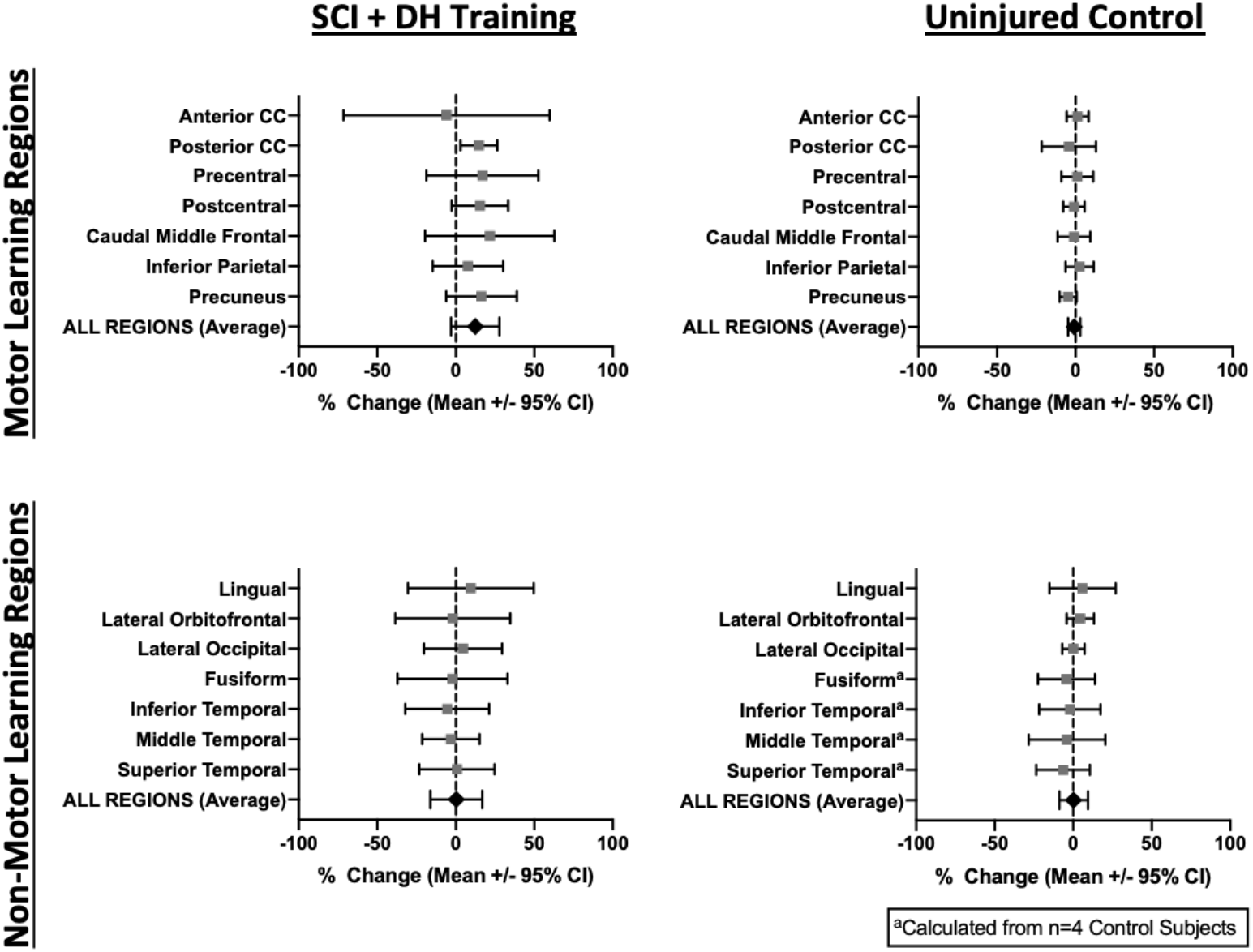
*Estimation-based approach shows specificity of brain MWF changes*. Plots of the mean % changes ± 95% confidence interval shows a specific pattern of MWF increase in motor learning regions of individuals with SCI who underwent downhill training (rightward shift). Within downhill-trained individuals, non-motor learning regions had no change in MWF. Uninjured controls also showed no change in MWF in motor learning and non-motor learning regions.

A spatially-distinct pattern also emerged in individuals with SCI as noted by a rightward shift in MWF CI in the ventral funiculi but a leftward shift in the dorsal columns. No pattern was present in the spinal cord of uninjured control participants (Fig. 9).

**Figure 9.**
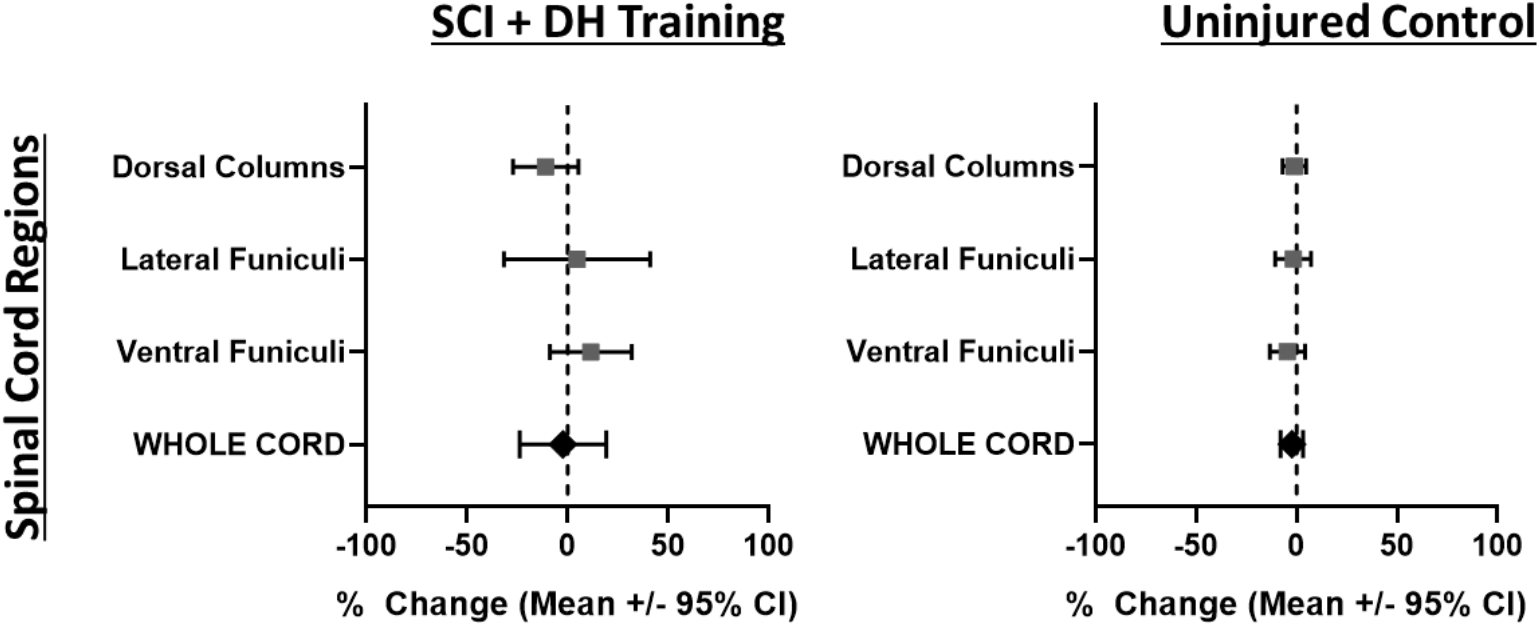
*Estimation-based approach shows specificity of spinal cord MWF changes*. Plots of the mean % changes ± 95% confidence interval shows a spatially-distinct pattern of MWF changes in the spinal cord after training.

## Discussion

Our results provide the first evidence of training-induced white matter plasticity in the brain and spinal cord of individuals with chronic, incomplete SCI. Importantly, MWF increases occurred in CNS regions primarily associated with motor learning and descending motor control. These findings extend previous rodent and human studies that show learning-induced white matter plasticity after complex motor task training (4-10). Here, we provide the first evidence of myelin-specific changes in the brain following a complex locomotor learning task.

Downhill training was predicted to spark an adaptive myelin response via release of neuroactive molecules, such as glutamate, along neurally-activated axons (12-22, 24-26). Adaptive myelination in response to neurotransmitters relies on differentiation of new oligodendrocytes (113). In rodents with SCI, some oligodendrocyte progenitor cell proliferation and differentiation occurs at the injury site (114-122) but whether rehabilitation training produces further gains is unknown. There are opportunities for new myelin to be added in response to training. First, gaps in myelin have been identified in the normal CNS which differs from the classic view of regularly spaced myelin nodes extending the full length of the axon (123, 124). Second, SCI reduces myelin below the injury as measured by MWF in rodents (44). Adding new myelin at these gaps and in areas of decreased myelin will speed up neural signaling and contribute to synaptic strengthening, a key component of motor learning (125).

Brain regions that responded to eccentric training occurred specifically in motor learning areas (i.e. white matter underlying primary motor/sensory, motor planning, and visuomotor association cortices). These motor learning regions are preferentially activated by and receive greater afferent feedback during eccentric contractions (67-71, 126). Increased MWF in the caudal middle frontal region after training in two out of three individuals with SCI highlights the cognitive demand on task performance and is indicative of early motor learning (127, 128). Indeed, individuals with SCI report having to consciously “think about” every aspect of every step and depend on vision when walking (129, 130). Visuomotor association areas, particularly the posterior corpus callosum, showed the most consistent MWF increases across individuals with SCI. These were unexpected changes given that downhill training avoided the use of mirrors and downward gaze. Perhaps the increased myelin in visual regions reflects motor learning effects off the treadmill during daily walking. Interestingly, virtual reality training for the lower extremities after SCI resulted in a different pattern of white matter changes, supporting that adaptive myelin plasticity is highly task specific (131). Taken together, our data support and extend past work highlighting the increased role of the brain in controlling locomotion after SCI. It will be important for future studies to perform MWI at multiple time points before, during, and after training to better understand the timing of MWF change in various brain regions and determine whether increases are maintained over time.

In the spinal cord, one individual (SCI-01) had increased MWF in the ventral funiculi but decreased MWF in the dorsal column sensory tract that exceeded the MDC (Fig. 3). The other two participants with SCI demonstrated this pattern although to a lesser extent. The concept of a training-induced decrease in MWF is interesting, as thinning of the myelin sheath by perinodal astrocytes was recently identified as a mechanism to finely tune conduction velocity and spike-time arrival (132). That myelin thickness could be increased or decreased to alter neural circuit synchrony during motor learning raises numerous questions about the type, timing, and sequence of rehabilitation approaches to maximize plasticity and function after CNS trauma. For instance, locomotor training on a flat treadmill, which was a prerequisite for participation, is a well-established model for sensory-driven locomotion via central pattern generator activation (133-138). Meanwhile, axons in the ventral spinal cord are major targets for collateral sprouting from supraspinal axons as they attempt to bypass the lesion (139) and are closely related to locomotor control after SCI (140-142). As such, the observed tendency for a decrease in dorsal column MWF alongside a concomitant increase in ventral MWF observed here could suggest a training-induced shift from sensory driven locomotion to more integrated control with greater input from descending systems.

An important aspect of the increased MWF with eccentric training is that it occurred at such substantial SCI chronicity - more than 40 years in two individuals. Thus, the potential for locomotor-dependent myelin plasticity may persist indefinitely after SCI. Based on animal SCI models, we presume that this plasticity reflects new motor learning given that neither general activity nor performing a previously learned task induced white matter plasticity (8). Importantly, this implies that eccentric motor control may not be trained by conventional treadmill-based rehabilitation or by living with SCI. Future studies into the evolution of myelin plasticity after SCI and with various rehabilitation approaches will add clarity.

An obvious limitation of this study is the small sample size. Spine hardware, claustrophobia, and motion artifact are MRI-specific complications encountered here. In future studies, interference by hardware can be minimized by adjusting MRI collection parameters to enable greater participation in imaging. In acknowledgement of the low number, we used within-subject brain regions as negative controls and statistical approaches appropriate for low sample sizes (MDC, confidence intervals, nonparametrics). The concurrence across individuals and positive myelin responses in *a priori* brain regions offers some confidence that our findings here might generalize to larger groups. Consideration of MWF as a measure of adaptive myelin after SCI carries risks that part of the value reflects iron content (143). We believe these risks are very low since these sequala will have resolved in chronic SCI. Also, MWF values agree well with histopathologically identified myelin (30, 31, 38-45). Even so, MWF cannot differentiate between the mechanisms of myelin plasticity such as *de novo* myelination by newly formed oligodendrocytes, new myelin internodes, increased internode length, or increased myelin thickness. While a combination of these mechanisms is likely involved in training-induced myelin plasticity, elegant work conducted in mice (8, 26) along with our own preclinical work (data not shown) indicate that new myelin produced by newly differentiated oligodendrocytes play a role.

Overall, these data are the first to show white matter plasticity in individuals with SCI and in response to a locomotor intervention. Even though from a small sample, these data are compelling and suggest that myelin is modulated by experience decades after SCI. Consideration of white matter plasticity as a relevant parameter in recovery of function is warranted. Future studies that establish the rate and extent of adaptive myelination in SCI are necessary to maximize recovery potential.

## Data Availability

The data that support the findings of this study are available from the corresponding author upon reasonable request.

## Acknowledgements

This work was funded by the National Institutes of Health [F31NS096921 (TDF), R21HD082808 (DMB)] and the Craig H. Neilsen Foundation [316282 (DMB)]. TDF was also supported by a Promotion of Doctoral Studies Level II Scholarship from the Foundation for Physical Therapy Research.

## Author Contributions

TDF, DMB, LAB, and DMM designed the study. TDF and DMB led downhill training sessions for individuals with SCI. TDF, HTN, PS, and MVK collected the magnetic resonance imaging data. TDF, BL, and HL processed the data. TDF, BL, HL, KRL, JLKK, LAB, and DMB analyzed and interpreted the data. TDF wrote the first and final drafts of the manuscript and prepared the figures, under the guidance of DMB. DMB wrote portions of the introduction and discussion. All authors edited the manuscript and approved the final version.

